# Disinfecting handheld electronic devices with UV-C in a healthcare setting

**DOI:** 10.1101/2020.04.01.20048496

**Authors:** Suzan Cremers-Pijpers, Carsten van Rossum, Heiman Wertheim, Alma Tostmann, Joost Hopman

**Affiliations:** Hygiene and Infection Control Unit, Department of Medical Microbiology, Radboudumc Center for Infectious Diseases, Radboud University Medical Centre, Nijmegen, The Netherlands

## Abstract

Handheld Electronic Devices (HEDs) play a central role in the current hospital environment. However, HEDs can be a potential vehicle for transmitting (pathogenic) microorganisms. We conducted a study to assess whether disinfection with UV-C light is successful in disinfecting three different handheld electronic devices in a clinical operational setting. More than half of the baseline measurements were moderately (>10CFU) or highly (>50 CFU) contaminated. Post-disinfection the CFU was 0 in 87% of measurements. We conclude that the UV-Smart D25 can successfully be used to disinfect non-critical handheld electronic devices in the clinical healthcare.

## Introduction

Proper cleaning and disinfection of non-critical devices in healthcare is essential for safe patient care. Current disinfection protocols recommend regular disinfection with disinfection wipes. If disinfection is not performed on at least a daily basis -and ideally after every use-, the bacterial burden is no different compared to ‘control’ devices that are not disinfected.^1^ Disinfection wipes are often based on alcohol or other compounds such as Quaternary Ammonium Compounds. They are however not recommended by manufacturers of handheld electronic devices (HEDs).

Handheld Electronic Devices (HEDs) play a central role in the current hospital environment, providing an accessible and portable method for delivering personalized healthcare.^2^ However, HEDs have also been shown to be a potential vehicle of transmission of (pathogenic) microorganisms. It has been described that up to 96% of healthcare workers (HCWs) do not regularly clean or disinfect their device and pathogenic microorganisms have been found on 9-27% of these devices.^3-5^

Novel disinfection methods for HEDs include the use of UV-C light, which has strong disinfecting properties.^6,7^ Additionally, UV-light has been shown to reduce bacterial load on HEDs within one minute to a minor fraction of the original load.^5^ Although the disinfection properties of the UV-Smart D25 has been shown in experimental settings, the effect in a clinical operational setting needs to be explored. We therefore conducted a study to assess whether the UV-Smart D25 is able to successfully disinfect three different handheld electronic devices in a clinical operational setting.

## Methods

### Study design

The UV-Smart D25 is a mobile UV-C device developed by UV-Smart that can disinfect mobile equipment and non-critical electronic devices in a healthcare setting.^8^ The UV-Smart D25 was piloted at the department of Internal Medicine and the Department of Orthopaedics of the Radboud university medical centre in Nijmegen, The Netherlands.

The effect of the UV Smart D25 was tested on surfaces of three different HEDs that are frequently used in our hospital by a wide range of healthcare professionals: a regular smartphone, a DECT phone (Digital European Cordless Telecommunications), and the Sensor ViSi Mobile Sotera touchscreen. The effect on bacterial contamination of these devices was assessed by counting the colony forming units (CFUs) on RODAC plates (Figure 1).

**Figure 1:**
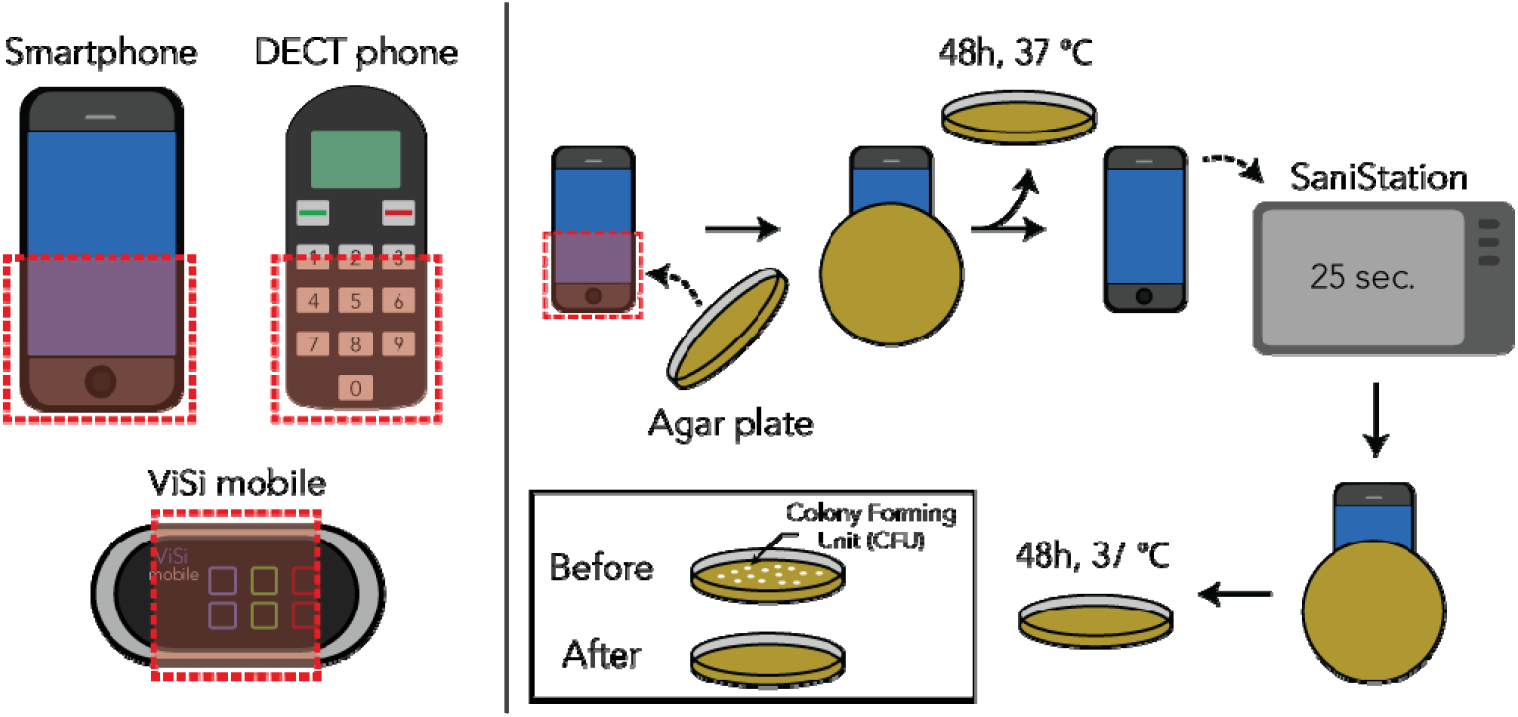
Schematic representation of the experimental set-up. Left: The areas of interest are indicated in red for the 3 unique devices: a smartphone (N=100 baseline and post-disinfection), DECT phone (N=200) and ViSi Mobile (N=100), respectively. Right: The number of colony forming units (CFUs) of a device at baseline and post disinfection using the UV-Smart D25 were measured using a total count RODAC plates.

The UV-Smart D25 was positioned at one nursing station and one nurse meeting room. They were not placed in or near patient care areas. Dedicated HCWs on the departments and an IPC trainee were instructed by an IPC professional how to use the UV-Smart D25, using an user’s manual and a short video created by the department of infection prevention and the manufacturer.

### Data collection

In December 2018 and January 2019, we took 100 samples from a Sensor ViSi Mobile Sotera touchscreen and 100 samples from a DECT Phone used by nurses at the Department of Internal Medicine. In the same time period, we took 100 samples from smartphones used by nurses and doctors and 100 samples from DECT Phones used by nurses at the Department of Orthopaedics. From each device, both baseline and a post-disinfection sample was taken, resulting in a total of 800 measurements (400 baseline and 400 post-disinfection).

### Data analysis

The CFU counts per RODAC plate were described as a median (interquartile range, IQR) due to the expected skewed distribution of these values. Median baseline CFU counts were compared between the different devices using a Mann-Whitney U test. Median baseline and post-disinfection CFU counts were compared using a Wilcoxon signed rank test for each of the three devices separately.

We discarded RODAC/CFU counts for both the baseline and post-disinfection test if one the test results could not be used due to contamination or other errors. The following thresholds for contamination were used: low level of contamination (<10 CFU/RODAC), moderate contamination (10-50 CFU/RODAC) and high level of contamination (≥50 CFU/RODAC).

## Results

In total we conducted 800 measurements, however we had to exclude 2 smartphone measurements, 6 DECT phone measurements and 4 ViSi Mobile measurements, resulting in a total of 98, 194 and 96 valid results for the smartphones, DECT phones and ViSi Mobiles touchscreens, respectively.

The baseline measurements showed a ‘high level of contamination’ (≥50 CFU) for 19% of the smartphone measurements, 10% of the DECT phone measurements and 31% of the ViSi Mobile touch screen measurements. The median (IQR) baseline CFU counts were 12 CFU/RODAC (12-44) for smartphones, 12 CFU/RODAC (5-27) for the DECT phones and 26 CFU/RODAC (14-60) for the ViSi Mobile touchscreens (Figure 2). Compared to the DECT phones, the CFU counts were higher on both smartphones (*P*<0.001) and ViSi Mobile touchscreen (*P* <0.001). There was no statistically significant difference in median CFU count between the smartphones and ViSi Mobiles (*P*=0.20).

**Figure 2.**
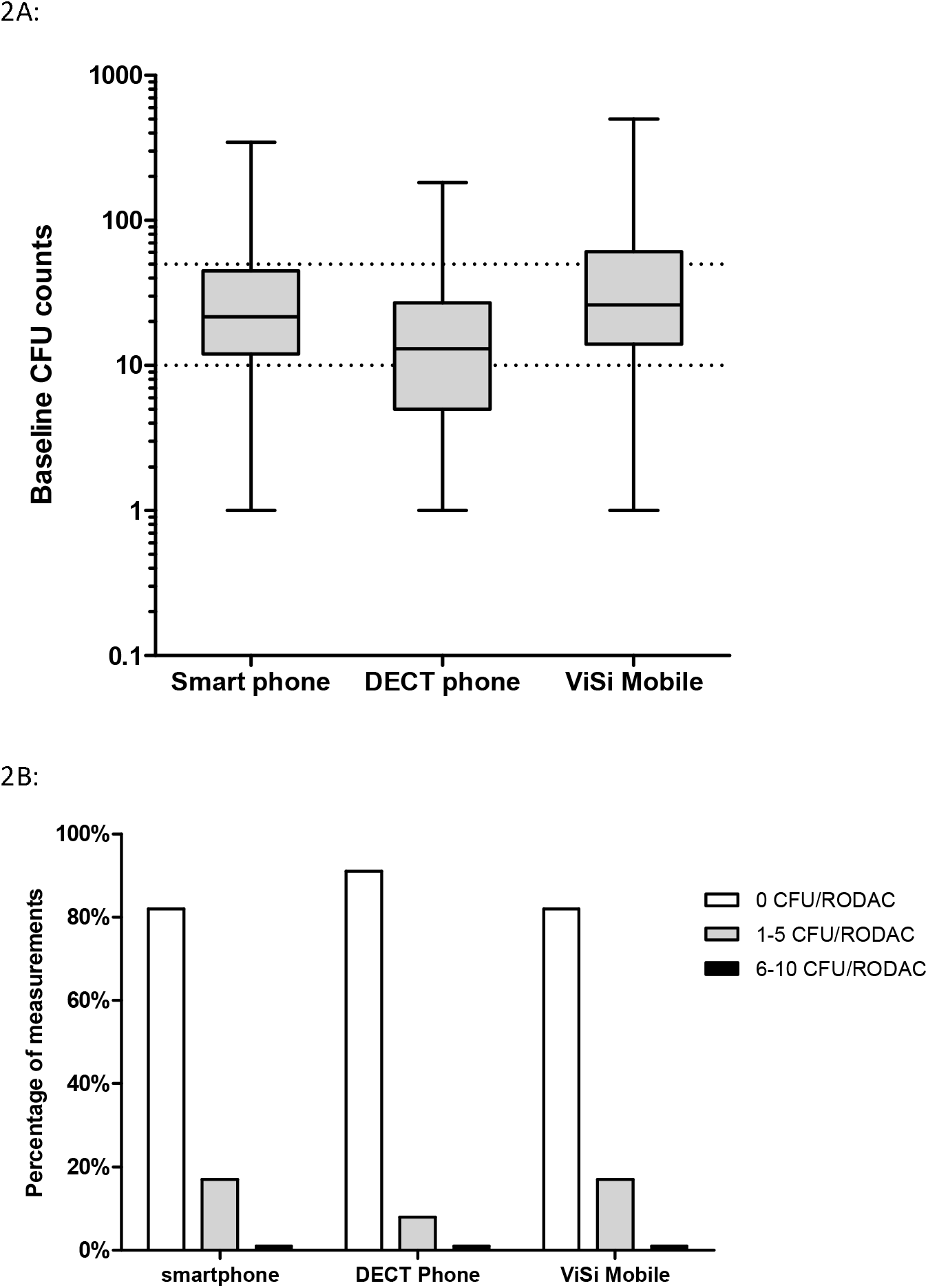
2A. Baseline CFU results for the smartphones (N=98), the DECT phones (N=194) and the ViSi Mobile touch screens (N=96). Dotted lines indicate the thresholds for low (<10 CFU) and high (>50 CFU) contamination level. 2B. Post-disinfection CFU/RODAC counts for the measurements on the smartphones (N=98), DECT phones (N=194) and ViSi Mobile touchscreens (N=96)

### Post-disinfection CFU counts

After UV-C disinfection, overall 87% of measurements had a complete reduction to 0 CFU. For the three devices, 82% of smartphone, 91% of the DECT phone and 82% of the ViSi Mobile touchscreen measurements were 0 CFU. This corresponded to a median (IQR) post-disinfection CFU counts of 0 CFU/RODAC (0-0) for all three devices. The CFU counts was only 1-2 CFU/RODAC in 12% of post-disinfection measurements, and the highest post-disinfection CFU count was 7 CFU/RODAC.

The comparison between baseline and post-disinfection CFU counts showed a statistically significant decrease for all three devices (P<0.001 for smartphones, DECT phones and ViSi Mobile touchscreens). The overall mean CFU reduction was 97.9%.

## Discussion

This study shows that in a clinical setting, the UV-Smart D25 reduces the bacterial contamination of smartphones, DECT phones and ViSi Mobile touchscreens to 0 CFU in 87% of measurements. This can be considered a large effect, especially in a context where the average handheld electronic device is contaminated, as was shown by our baseline measurements.

The post UV-C reduction in bacterial load was to be expected, since the effectivity of UV-C light for disinfecting smartphones and tablets has been shown to be more effective than other sanitization methods. ^6,9^ Others also found (almost) complete reductions in bacterial loads in cycles of 35-60 seconds, whereas Huffman et al. stated that no bacterial growth was observed after having inoculated inactive phones and ID-badges for 48 hours with known amounts of bacteria and subsequent disinfection.^5,9^

The remaining bacterial load that was observed in this study may be explained by shadows formed by the HEDs. These are places not reached by the UV-light. For example, Wallace et al. -who also concluded that UV-C light was capable of reducing MRSA, C. difficile and norovirus load in 30 to 60 seconds-observed that bacterial load reduction plateaus before maximum intensity was reached.^10^ They attributed this to the high inoculum concentration, whereas the bacteria on the surface were neutralized and created a shadow for the (surviving) bacteria underneath. This shadowing principle can also be applied to HEDs in the UV-Smart D25. However, these remaining low bacterial counts should be viewed in the context of high baseline levels of contamination.

A limitations of this study was that we only included two departments. It would be interesting to repeat this study on different departments in a multi-centre study. We have only tested three specific handheld electronic devices. Tablets have not been assessed, however, and are becoming more apparent in patient rooms as patient-bound devices. Other noncritical devices to be considered include stethoscopes, blood pressure machines, glucose machines or scissors.

A challenge for implementation of a UV Smart D25 would be to decide on a suitable and feasible frequency of use. Even though bacterial loads get below the detection limit directly after disinfection by UV-C, recontamination via hands and other -often contaminated-hospital environments occurs relatively quickly, returning to pre-disinfection levels within 48 hours.^9^ These studies suggest that a daily or twice daily disinfecting round with UV-C should result in a consistent low bacterial load. Upon implementation, placement of the UV-Smart D25 should be considered carefully. They may be too large to be placed in the patient room and they are vulnerable in the corridor. A suitable option is the nursing station or the coffee or lunchroom for HCWs, as we did in this pilot. The UV Smart is a valuable addition to the disinfection options in a healthcare setting, provided proper instructions and clear protocols are developed. The 25-seconds ‘waiting time’ provides a good opportunity for hand disinfection, as was instructed during this pilot.

Our study focused on the reduction of bacterial load on the HEDs. In the context of the current SARS-CoV2 pandemic, it would be worthwhile to investigate the effect of UV-C on SARS-COV-2 on the surface of HEDs and non-critical devices and medical N95 respirators.

In conclusion, this study showed that the UV-Smart D25 can successfully be used to disinfect non-critical handheld electronic devices which are used in the clinical healthcare. However, further applied research is required on the best method to implement the machine in clinical practice to ensure ease of use, high compliance and effective disinfection cycles.

## Data Availability

For requesting any data gathered during the measurements, please contact the corresponding author.

## Acknowledgements

We thank our colleagues at the Department of Internal Medicine and the Department of Orthopaedics for their cooperation. We have no conflicts of interest. The UV Smart D25 was made available for this pilot project by UV Smart Company, Delft, The Netherlands.

## Declaration of interest

Declarations of interest: none

## Source of funding

This research did not receive any specific grant from funding agencies in the public, commercial, or not-for-profit sectors. The UV Smart D25 machine from UV Smart was kindly provided by the company.

## Notes

### Competing Interest Statement

The authors have declared no competing interest.

### Funding Statement

No external funding was recieved.

## References

1. Koscova J, Hurnikova Z, Pistl J. Degree of Bacterial Contamination of Mobile Phone and Computer Keyboard Surfaces and Efficacy of Disinfection with Chlorhexidine Digluconate and Triclosan to Its Reduction. Int J Environ Res Public Health 2018;15.

2. Mickan S, Tilson JK, Atherton H, Roberts NW, Heneghan C. Evidence of effectiveness of health care professionals using handheld computers: a scoping review of systematic reviews. J Med Internet Res 2013;15:e212.

3. Brady RR, Verran J, Damani NN, Gibb AP. Review of mobile communication devices as potential reservoirs of nosocomial pathogens. J Hosp Infect 2009;71:295–300.

4. Khan A, Rao A, Reyes-Sacin C, et al. Use of portable electronic devices in a hospital setting and their potential for bacterial colonization. Am J Infect Control 2015;43:286–8.

5. Huffman S, Webb C, Spina SP. Investigation into the cleaning methods of smartphones and wearables from infectious contamination in a patient care environment (I-SWIPE). Am J Infect Control 2019.

6. Lieberman MT, Madden CM, Ma EJ, Fox JG. Evaluation of 6 Methods for Aerobic Bacterial Sanitization of Smartphones. J Am Assoc Lab Anim Sci 2018;57:24–9.

7. Mathew JI, Cadnum JL, Sankar T, Jencson AL, Kundrapu S, Donskey CJ. Evaluation of an enclosed ultraviolet-C radiation device for decontamination of mobile handheld devices. Am J Infect Control 2016;44:724–6.

8. https://www.uvsmart.nl/. 2020.

9. Muzslay M, Yui S, Ali S, Wilson APR. Ultraviolet-C decontamination of hand-held tablet devices in the healthcare environment using the Codonics D6000 disinfection system. J Hosp Infect 2018;100:e60–e3.

10. Wallace RL, Ouellette M, Jean J. Effect of UV-C light or hydrogen peroxide wipes on the inactivation of methicillin-resistant Staphylococcus aureus, Clostridium difficile spores and norovirus surrogate. J Appl Microbiol 2019;127:586–97.

